# An experimental trial of recombinant human interferon alpha nasal drops to prevent COVID-19 in medical staff in an epidemic area

**DOI:** 10.1101/2020.04.11.20061473

**Authors:** Zhongji Meng, Tongyu Wang, Li Chen, Xinhe Chen, Longti Li, Xueqin Qin, Hai Li, Jie Luo

## Abstract

**Objective:** To investigate the efficacy and safety of recombinant human interferon alpha (rhIFN-α) nasal drops in healthy medical staff to prevent coronavirus disease 2019 (COVID-19).

**Methods:** A prospective, open-label study was conducted in January 21, 2020at Taihe Hospital in Shiyan City, Hubei Province. Totally, 2944 medical staff members were recruited and allocated into low-risk group or high-risk group according to whether they were directly exposed to COVID-19 patients. Participants in the low-risk group received rhIFN-α nasal drops (2–3 drops/nostril/time, 4 times/day) for 28 days with first-level protection; those in the high-risk group received identical rhIFN-α nasal drops combined with thymosin-α1 (1.6 mg, hypodermic injection, once a week) along with secondary-level or third-level protection. The primary outcome was new-onset COVID-19 over 28 days. The secondary outcome was new-onset fever or respiratory symptoms but with negative pulmonary images. The results were compared with new-onset COVID-19 in medical staff in Hubei Province (including Wuhan) during the same period. Adverse reactions to interferon nasal drops were also observed.

**Results:** Among the 2944 subjects in our study, 2415 were included in the low-risk group, including 997 doctors and 1418 nurses with average ages of 37.38 and 33.56 years, respectively; 529 were included in the high-risk group, including 122 doctors and 407 nurses with average ages of 35.24 and 32.16 years, respectively. The 28-day incidence of COVID-19 was zero in both the high and low-risk groups. The 28-day incidence of new-onset clinical symptoms with negative images for pneumonia was also zero in both the high and low-risk groups. As control, a total of 2035 medical personnel with confirmed COVID-19 from the same area (Hubei Province) was observed between January 21 to February 23, 2020. No serious adverse events were observed in our trial during the intervention period.

**Conclusion:** In this investigator-initiated open-label study, we observed that rhIFN-α nasal drops may effectively prevent COVID-19 in medical staff, as an enhancement protection on the basis of standard physical isolation. Our results also indicate that rhIFN-α nasal drops have potential promise for protecting susceptible healthy people during the coronavirus pandemic.

## Introduction

Coronavirus disease 2019 (COVID-19) caused by the severe acute respiratory syndrome coronavirus 2 (SARS-CoV-2) initially reported in Wuhan City, Hubei Province, China in December 2019 and January 2020. It is a severe respiratory disease associated with high mortality in critical cases ^[1, 2]^. As of April 15, 2020, there have been more than 1.9 million individuals affected and nearly 12300 deaths over the world ^[3]^. Before government intervention and medical assistance from other provinces of China, health care workers in Wuhan and some other cities in Hubei province faced overwhelming work intensity and high risk of nosocomial infection, accounting for 90% infected medical staff in our country ^[4, 5]^.

SARS-CoV-2 is primarily transmitted through the respiratory tract and direct contact. Air and contact isolation are pivotal to reduce the spread of the virus, COVID-19 patients and those infected with SARS-CoV-2 should be isolated in single-room, with constant air circulation, and disinfection of the air and environmental items ^[6]^. A mask (N95) can block 95% of the virus from entering the respiratory tract. Strict implementation of hand hygiene can prevent contact transmission. Due to the intense outbreak of COVID-19 over a short period in early Hubei, especially in Wuhan, the isolated wards of medical institutions are unable to accommodate the admission of a large number of patients. Many ordinary wards have been temporarily converted into isolated wards, which fail to meet the “two-zone, three-passage” standard. Furthermore, in the early stage of the epidemic, protective supplies such as N95 masks were scarce, a part of medical staff lacked awareness of self-protection, failed to standardize mask-wearing and hand hygiene, leading to a large number of infections among medical staff. Therefore, it is urgent to assure occupational safety and health in medical staff during early stage of disease transmission, when there was neither effective drug nor vaccine.

Medical personnel, especially front-line medical staff involved in fever clinics and isolation wards, have long been engaged in the diagnosis and treatment of COVID-19. Consequently, they have been at high risk of SARS-CoV-2 infection for a long time. Even if the standard second-level protections, such as a disposable round cap, gown, protective clothing, N95 mask and surgical mask, and double gloves, are implemented and enhanced second-level protections (e.g., the use of a protective screen or positive pressure breathing mask during airway operations) are put into use when necessary, these measures do not completely protect people from infection with SARS-CoV-2.

In cases of SARS-CoV-2 invasion of the mouth and nose, immune intervention strategies to increase the local or systemic immunity may build up the body’s resistance to the virus and cover the shortage of physical protection. Interferon (IFN) is by far the most widely used antiviral biological drug discovered in 1957 ^[7]^. It is an important cytokine that regulates cell functions and block virus particles replication, and has effect on both DNA and RNA viruses, it may reduce the amount of virus, lead to restriction and even clearance of the virus. For general viral infection, IFN can shorten the course of the disease ^[8]^. Moreover, IFN nasal drops maintain a high IFN-α concentration in the nasal mucosa, which can inhibit virus accumulated in the nasal mucosal epithelium and prevent from break through the mucosal immune barrier. According to the latest results from the P4 laboratory of the First Affiliated Hospital of Zhejiang University, China, SARS-CoV-2 can reduce the ability of host cells to suppress viruses by reducing endogenous IFN production. Therefore, the use of exogenous IFN in early stage may be important.

A trial on the preventive effect of recombinant human IFN-α nasal drops against COVID-19 in medical staff has been carried out, and the results will be presented in the present study.

## Methods

### Study population

Since January 21, 2020, 2944 official medical staff, including doctors and nurses from Taihe Hospital, Shiyan City, Hubei Province, have been included in the study. Among them, 2415 medical staff who were working in non-isolated wards and non-fever clinics were categorized into the low-risk exposure group. They were not in direct contact with SARS-CoV-2 infected patients. In contrast, 529 medical staff who were working in isolated wards and fever clinics for diagnosis and treatment of COVID-19 were categorized into the high-risk exposure group. The personnel in this group were in direct contact with SARS-CoV-2 infected patients. Baseline information is shown in Table 1. The study was approved by the Medical Ethics Committee of Taihe Hospital, Shiyan City. This study has been registered in the clinicaltrials.gov and the registration number is NCT04320238.

**Table 1.**
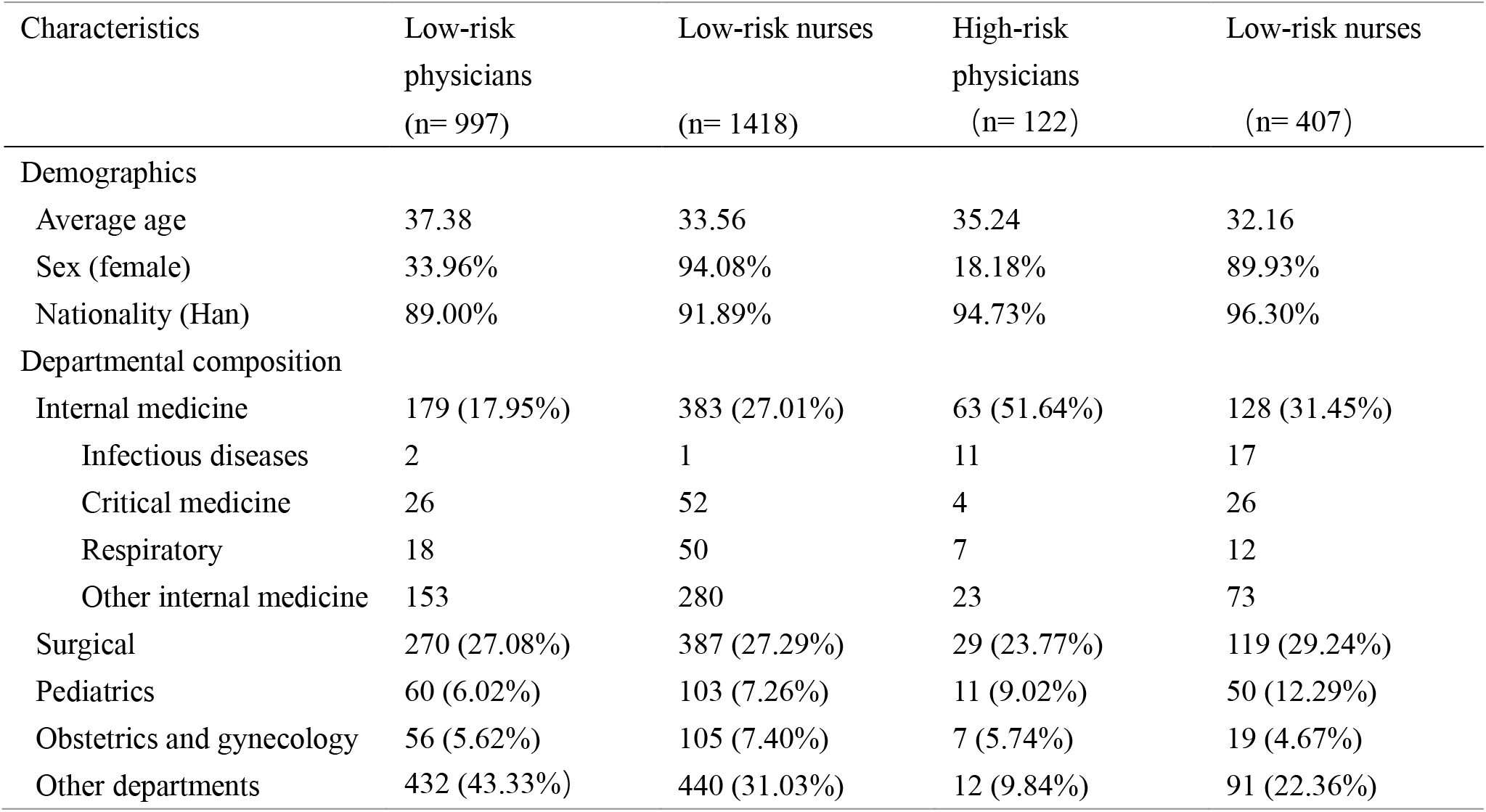
Baseline characteristics of the low-risk group that received interferon nasal drops and the high-risk group that received interferon nasal drops combined with thymosin-α1 injection at Taihe Hospital in Shiyan City, Hubei Province

### Diagnostic criteria

Diagnosis was made according to the “Diagnosis and Treatment Protocol for Novel Coronavirus Pneumonia” issued by the General Office of the National Health Commission of the People’s Republic of China ^[9]^, as follows:

SARS-CoV-2 infection: Positive pathogenic test (nucleic acid or specific antibody), including:

a. Asymptomatic infection: The total number of white blood cells was normal or decreased in the early stage of onset and the lymphocyte count was normal or decreased. There was no fever and/or respiratory symptoms and no imaging characteristics of pneumonia.
b. Asymptomatic non-pneumonia COVID-19: Normal or decreased white blood cells in the early stages of onset, lymphocyte counts were normal or decreased, and fever and/or respiratory symptoms were observed. There were no imaging characteristics of pneumonia.
c. COVID-19 pneumonia confirmed cases: In the early stage of onset, the total number of white blood cells was normal or decreased, and the lymphocyte counts were normal or decreased. Fever and/or respiratory symptoms were present. In addition, image characteristics of novel coronavirus pneumonia were found, for example, in the early stage, multiple small patchy shadows and interstitial changes were evident in the extrapulmonary zone. Furthermore, multiple ground glass infiltrations and infiltrates could be detected in both lungs. In severe cases, pulmonary consolidation occurred. Pleural effusion was rarely seen.

### Inclusion and exclusion criteria

Inclusion criteria: Official members of Taihe Hospital medical staff.

Exclusion criteria: Participants who met the following conditions were excluded: pregnant women, those with severe chronic illnesses who were unable to participate in normal health care work, those with acute fever (>37.2 centigrade) and/or respiratory symptoms who were unwilling to receive IFN-α nasal drops or thymosin-α1.

### Intervention measures

Infection prevention and control standards

First-level protections: Wearing work clothes, disposable round caps and disposable medical surgical masks (replaced every 4 hours) and strictly implementing hand hygiene.

Secondary-level protection: Wearing work clothes, protective clothing, medical protective masks, goggles/protective screens, disposable round caps and gloves, double gloves if necessary, shoe covers/boot covers and work shoes/rubber boots.

Third-level protections: Wearing a comprehensive protective mask and double gloves in addition to the secondary-level protection.

#### Interventions

Low-risk group intervention: In addition to the first-level protections, rhIFN-α nasal drops (Beijing Tri-Prime Gene Pharmaceutical Co., Ltd., China, IFN-α 1b, 3000 µ/ml, in-hospital preparation) were administered at a dosage of 2-3 drops/nostril/time, four times/day. The intervention duration was 28 days.

High-risk group intervention: In addition to the secondary-level protections (and third-level protections, if necessary), rhIFN-α nasal drops were administered as indicated above; additionally, thymosin-α-1 (Chengdu Shengnuo Biotech Co., Ltd., China, 1.6 mg/tube) was injected subcutaneously at a dosage of 1.6 mg, once a week. The intervention duration was 28 days.

All participants were closely followed for 28 days during the intervention. For both groups, routine etiological tests (including pharyngeal swab nucleic acid and serum antibody tests) would be performed if fever and/or respiratory symptoms were observed.

### Primary and secondary outcomes

The primary outcome was the development of COVID-19 pneumonia by the 28th day after the preventive drug intervention. The secondary outcome was the new-onset clinical symptoms of COVID-19 without image findings of pneumonia. Reported cases of COVID-19 pneumonia in Wuhan, in Hubei Province (other than Wuhan), and among nationwide medical staff reported in a contemporary study were used as control. Additionally, the adverse reactions after the use of rhIFN-α nasal drops was observed.

## Results

### Efficacy of rhIFN-α nasal drops and in combination with thymosin-α1 for preventing COVID-19

In the low-risk exposure group, 2415 medical staff members were treated with rhIFN-α nasal drops alone for 28 days. No new cases of COVID-19 pneumonia were confirmed during follow-up. New pulmonary image of viral pneumonia-like features was negative, and zero staff members developed fever/respiratory symptoms. As of March 6, no newly confirmed cases of COVID-19 pneumonia were found during the follow-up.

In the high-risk exposure group, 529 medical staff members received rhIFN-α nasal drops combined with thymosin-α1 for 28 days. During follow-up, no new cases of COVID-19 pneumonia were diagnosed. The medical staff were negative with the onset of new pulmonary image of viral pneumonia-like features, with zero confirmed cases of fever/respiratory symptoms. As of March 6, no new confirmed case of COVID-19 pneumonia was found during the follow-up.

The control group was drawn from another study on medical staff diagnosed with COVID-19 pneumonia nationwide and in Wuhan from January 1 to February 11, 2020^[5]^. Chinese medical staff diagnosed with COVID-19 pneumonia as reported by the China-World Health Organization joint inspection expert group as of February 23 were also included in the control group. In 422 medical institutions providing COVID-19 diagnosis and treatment service, a total of 3387 medical staff were etiologically diagnosed (1716 cases), clinically diagnosed (1070 cases) or suspected (157 cases) as COVID-19 pneumonia, among them 3062 cases came from Hubei (Table 2).

**Table 2:** New-onset COVID-19 pneumonia of medical staff treated with interferon and interferon nasal drops combined with thymosin-α1 from January 25 to February 22, 2019 in Taihe Hospital, Shiyan, Hubei, and in other regions during the same period. -Confirmed cases, severe cases, death cases

| Time | Medical staff at low-risk group in Taihe Hospital |  |  | Medical staff at high-risk group in Taihe Hospital |  |  | Wuhan medical staff § |  |  | Hubei (except Wuhan) medical staff § |  |  | Nationwide (except Hubei) medical staff § |  |  |
| --- | --- | --- | --- | --- | --- | --- | --- | --- | --- | --- | --- | --- | --- | --- | --- |
|  | Confir med cases | Severe (%) | Deaths (%) | Confir med cases | Severe (%) | Deaths (%) | Confirm ed cases | Severe (%) | Deaths (%) | Confirm ed cases | Severe (%) | Deaths (%) | Confirm ed cases | Severe (%) | Deaths (%) |
| Before December 31, 2019 | 0 | 0 | 0 | 0 | 0 | 0 | 0 | 0 | 0 | 0 | 0 | 0 | 0 | 0 | 0 |
| January 1-10, 2020 | / | / | / | / | / | / | / | 7 (38.9) | 1 (5.6) | 1 | 1 (100) | 0 | 1 | 1 (100) | 0 |
| January 11-20, 2020 | / | / | / | / | / | / | / | 52 (22.3) | 1 (0.4) | 48 | 8 (16.7) | 0 | 29 | 1 (3.4) | 0 |
| January 21-31, 2020 | 0 | 0 | 0 | 0 | 0 | 0 | 0 | 110 (16.8) | 0 | 250 | 29 (11.6) | 2 (0.8) | 130 | 10 (7.7) | 0 |
| February 1-11, 2020 | 0 | 0 | 0 | 0 | 0 | 0 | 0 | 22 (12.7) | 1 (0.6) | 95 | 3 (3.2) | 0 | 54 | 3 (5.6) | 0 |
| February 12-22, 2020 | 0 | 0 | 0 | 0 | 0 | 0 | 0 | unknown | unknown | 80* | unknown | unknown | 37* | unknown | unknown |

§The data for Wuhan, Hubei, and the entire country come from the literature and the China-World Health Organization joint inspection expert group as of February 23. (3387 medical personnel from 476 medical institutions across the country have been infected with COVID-19 pneumonia; including 2055 confirmed cases, 1070 clinically diagnosed cases and 157 suspected cases. More than 90% of infected medical staff were from Hubei.)

### Safety of rhIFN-α nasal drops

According to previous studies, common adverse reactions of IFN spray/nasal drops include flu-like symptoms; slight local irritation such as burning pain and itching; allergic reactions such as rash, nausea, chest distress, palpitation and flushing were rare. No flu-like symptoms were observed among nearly 2,000 participants in our study. A few of participants experienced transient irritation of nasal mucosa such as itching, which relieved without interrupting the intervention.

## Discussion

This study preliminarily explores the preventive effect of rhIFN-α nasal drops against SARS-CoV-2 infection in medical staff over a period of 28 days. The study shows that if standard first- and second-level protections are strictly implemented, the use of IFN-α nasal drops may effectively prevent medical staff at low exposure level (i.e., those not directly exposed to COVID-19) from COVID-19 pneumonia. In addition, IFN-α nasal drops combined with weekly thymosin-α1 subcutaneous injection may help to prevent medical staff at high exposure level (i.e., those who are in direct contact with COVID-19 patients) from developing the disease.

COVID-19 is highly contagious and presents a long-lasting epidemic trend. COVID-19 has now progressively expanded to more than 200 countries and the global pandemic trend is apparent ^[10]^. At the beginning of transmission, health care workers were vulnerable populations with substantially high risk of nosocomial infection. Recently, a large number of medical staff infection in Italy and other countries have also been reported ^[11, 12]^. In addition, the study suggests that compared to those who were infected with SARS-CoV-2 before January 23, recently infected patients displayed more subtle symptoms, while the infectivity has not changed significantly, indicating that SARS-CoV-2 tends to gradually evolve into a low-virulence, highly infectious influenza-like virus ^[13]^. Therefore, experts predict that epidemics of the virus SARS-CoV-2 may recur every autumn and winter, and the virus will coexist with human beings for a long time. For this reason, clinicians and public health scholars currently believe that a broad-spectrum antiviral drug that inhibits coronavirus is essential to prevent recurrent epidemics and mutations both before and after a vaccine.

The keys to preventing and controlling COVID-19 are controlling the source of infection, cutting off the route of transmission, and protecting susceptible populations ^[14]^. In accordance with China’s “Diagnosis and Treatment Protocol for Novel Coronavirus Pneumonia (Fifth Edition)”, medical personnel working in outpatient departments with suspected or confirmed patients in the ward are equipped with second-level protection for the general diagnosis and treatment of patients. Medical staff involved in the treatment of suspected or confirmed patients using aerosol-generating procedures (such as tracheal intubation and related operations, cardiopulmonary resuscitation, bronchoscopy, sputum suction, throat swab sampling and the use of high-speed equipment (such as drilling, sawing, centrifuging, etc.), should use third-level protection. Medical staff in general wards should implement first-level protection and strictly follow standard hand hygiene procedures. Hand hygiene should be performed before putting on gloves and after removing gloves. Taihe Hospital in Shiyan City and other hospitals in Hubei Province have all followed the above-described prevention and control plan throughout the diagnosis and treatment of COVID-19. In addition to following these standard protections, the medical staff of Taihe Hospital in Shiyan City were given IFN-α nasal drops with or without subcutaneous thymosin-α1 according to their degree of exposure to COVID-19. Among the medical staff treated with this protocol, there were no COVID-19 cases reported over a 28-day observation period. In contrast, over the same period, over 100 cases of COVID-19 were confirmed among medical staff at more than 10 other hospitals in Hubei Province. The results suggest that IFN-α nasal drops, especially in combination with thymosin-α1, may improve the protection of medical staff against COVID-19 and are an effective supplement to physical protections.

Vaccines and prophylactic drugs are the two main approaches for protecting susceptible populations. Although many clinical trials of vaccines against COVID-19 have been launched, whether they are safe and can trigger immune responses is uncertain ^[15, 16]^. They might come too late to affect the first wave of this pandemic. Additionally, the existence of two subtypes of SARS-CoV-2 (the L and S subtypes) has been confirmed, and the possibility of new mutations cannot be ruled out ^[17]^. Therefore, experts have raised concerns about whether vaccines developed for early viral RNA sequences will be able to effectively protect against mutant viruses. Thus, there is an urgent need for drugs that can effectively prevent coronavirus infection in healthy people before a vaccine becomes available on the market and after the subsequent decline in the protection capability of early vaccines due to virus mutation.

IFN is a type of cytokine produced when a cell is stimulated by a viral infection or other IFN-inducing agent. It is a secreted protein (mainly a glycoprotein) that has many biological functions, including the regulation of innate and acquired immune responses after infection, in addition to a broad spectrum of antiviral, immune-regulating biological functions. IFN is divided into three types: I, II and III. Type I includes IFN-α and IFN-β; type II has only one subtype, IFN-γ; and type III includes IFN-λ 1 (IL-29), IFN-λ 2 (IL-28a), and IFN-λ (IL-28b). At present, the α subtype of type I IFN is generally used for the development of antiviral drugs because of its strong inhibition of viral replication. According to its amino acid sequence, IFN-α has more than 20 subtypes. Common IFN-αs that have been developed and marketed for antiviral therapy are IFN-α 1b, IFN-α 2b, IFN-α 2a, and IFN-ω. The antiviral mechanism of IFN is implemented by activating cell membrane adenylate cyclase on cell surface receptors while promoting the increase of adenylate cyclase, which activates the intracellular antiviral mechanism so that a group of antiviral substances, including anti-viral proteins and enzymes, can be generated. The generation of these substances has the effect of inhibiting virus replication and blocking virus spread.

Clinically, IFN-α has been used for a long time to prevent and treat the common cold and flu, especially in the early stage of epidemic. IFN-α is effective not only for preventing infection with influenza viruses, rhinoviruses but also coronaviruses in susceptible people ^[18]^. Since the severe acute respiratory syndrome (SARS) outbreak in 2003, the efficacy of IFN-α has attracted researchers’ attention. An in vitro test confirmed the efficacy of IFN-α against SARS-like coronavirus infection ^[19]^. Animal tests have confirmed that IFN-α nasal spray can effectively prevent or reduce coronavirus infection-related damage in monkeys ^[20, 21]^. In addition, a clinical study indicated that rhIFN-α 2b spray can reduce common respiratory virus infection to varying degrees with well safety in this study of 14,391 healthy people ^[22]^.

Evidence suggests that each type of IFN may play an important role in host defense and immunopathology, particularly at epithelial barriers. Recently, a study based on scRNA-seq datasets reveals that nasal goblet and ciliated cells highly express ACE2, receptor of SARS-CoV-2 invasion and TMRPSS2 viral entry-associated protease, implicating that these cells are the initial invasion sites and may play as virus reservoir for the disease transmission ^[23]^. Another article elaborated that IFN-α2 and IFN-γ drives ACE2 expression dose-dependently in primary human nasal epithelial cells. A dramatic reduction of virus replication after the addition of in vitro type I IFN was observed, suggesting that SARS-CoV-2 may inhibit subsequent upregulation of interferon stimulated genes (ISGs) ^[24]^. Thus, we hypothesis that exogenous addition of type I IFN in nasal epithelial cells may prevent virus invasion and transmission between individuals by creating a high local concentration of IFN, inhibiting virus replication in nasal goblet and ciliate cells, and reducing virus reservation.

Our study began on January 25, and there were no new confirmed cases of COVID-19 among 2944 medical staff members after 28 days of nasal drop intervention. The 2,415 medical staff members without direct exposure to COVID-19 were considered a healthy susceptible population, and the results confirm the effectiveness of IFN-α nasal drops for the prevention of COVID-19 in the general healthy population. The 529 medical staff members working in the isolation ward were also free of infections and confirmed cases, proving that the drug can strengthen the protection of medical staff in the isolation ward. These results indicate that for SARS-CoV-2, which is homologous to the SARS coronavirus, rhIFN-α nasal drops can effectively compensate for the deficiency of the physical barrier and improve the nonspecific antiviral effect against SARS-CoV-2 in susceptible populations, and the mechanisms of this effect are clear. In addition, it can reduce the vulnerability of SARS-CoV-2 resistance. Furthermore, the safety of intranasal IFN-α nasal drops has been effectively evaluated, and no serious adverse reactions or adverse events occurred in the present study.

This clinical study has several characteristics. Firstly, it is the first real-world study of an intervention using IFN-α nasal drops to prevent COVID-19 pneumonia in healthy susceptible people in the COVID-19 epidemic area. Secondly, this study began in the early COVID-19 outbreak in Hubei. Four weeks after the initiate of the study, the number of COVID-19 pneumonia cases in Hubei Province increased rapidly, while there were no new cases of COVID-19 pneumonia among the more than 2,000 participants in our tertiary hospital. This indicates that IFN-α has a promising preventive effect in people who are susceptible to the virus. Furthermore, this study divided the participants into low-risk group and a high-risk susceptibility group, single drug (IFN-α nasal drops) or combination drug (IFN-α nasal drops combined with thymosin-α1 subcutaneous injection) was administered respectively to the two groups with different risks of COVID-19. This provides insights and corresponding evidence related to the adoption of differentiated drug prevention methods for susceptible populations at different exposure levels.

Our study also has limitations. The study was not clustered, randomized designed with the control group from other epidemic area during the same period reported in the literature, rather than a strictly parallel, placebo-controlled group. Nevertheless, our study represents a real-world effort initiated by researchers in an emergent medical situation to prevent and control nosocomial infection in our medical staff. The results lay a good foundation for subsequent high-quality randomized, parallel, placebo-controlled studies. Secondly, SARS-CoV-2 nucleic acid tests and serum antibodies tests were not performed for the subjects in this study, so it was impossible to determine whether IFN-α has prevents infection with SARS-CoV-2; instead, the development of COVID-19 pneumonia was the main evaluation indicator. The main reason for using this outcome was that no COVID-19 diagnostic kits had been approved at the beginning of the study. Subsequent high-quality studies should use combined nucleic acid and serum antibody testing to diagnose SARS-CoV-2 infection.

In summary, in more than 2,000 health care workers at large tertiary hospitals who were at high or low risk of exposure to SARS-CoV-2 in Shiyan City, Hubei Province, a COVID-19 epidemic area, a real-world study of the use of IFN-α nasal drops with or without thymosin-α1 as a 28-day intervention shows that low-risk subjects treated with IFN-α nasal drops alone and high-risk subjects treated with IFN-α nasal drops combined with thymosin-α1 developed zero cases of COVID-19 pneumonia. We explore that rhIFN-α nasal drops can be used as an alternative drug to effectively prevent SARS-CoV-2 virus infection and protect healthy and susceptible people from coronavirus infection. During the COVID-19 epidemic, rhIFN-α nasal drops could play a complementary role along with vaccines. In the future, more large-scale clinical studies can be conducted to further verify the preventive effect of rhIFN-α nasal drops.

## Data Availability

The data used to support the findings of this study are included within the article.

